# Investigation of Dengue Fever Outbreak in a Rural Area of Islamabad, Pakistan: A Case Control Study

**DOI:** 10.1101/2021.10.01.21264329

**Authors:** Fawad Khalid Khan, Khurram Shahzad Akram, Ambreen Chaudhry, Mir Hassan Bullo, Zakir Hussain, Mirza Amir Baig, Amjad Mehmood, Zeeshan Iqbal Baig

## Abstract

**Background:** In the second week of October 2019, five suspected cases of dengue fever were reported from union council Sohan, Islamabad rural (population 45,747) to the health department, Islamabad Capital Territory (ICT). Outbreak investigation was conducted with the objectives to identify risk factors and recommend control measures.

**Methods:** Outbreak investigation was conducted from 17^th^ October to 25^th^ November 2019. A case was defined as, “fever and two or more of the following signs/symptoms; headache, retro-orbital pain, joint/bone pain, myalgia and petechial rash with NS1 test (Nonstructural Protein 1) positive during 8^th^ October to 25^th^ November 2019 among residents of Sohan”. Age and sex-matched controls were recruited from the same neighborhood. All cases were positive for NS1 antigen. Blood samples from five suspected cases were collected and tested for laboratory confirmation.

**Results:** A total of 547 households were surveyed and 85 cases were identified. The mean age was 34.4 years + 16.05 (range 3-71 years). The attack rate was 0.19% whereas the most affected age group was the 45-54 years (AR 0.43%). Males were predominantly affected (n=48 56.5%). Among all cases, 32% (n=27)) had stagnant water inside or around their houses (aOR 2.65, CI 1.20-5.83, *P=*0.005), 33% (n=28) were using mosquito repellent (aOR 0.35, CI 0.17-0.70, *P*<0.001), 31% (n=26) used indoor residual spray insecticide (aOR 0.48, CI 0.24-0.97, *P*=0.041), and 73% (n=62) used full protective clothing (aOR 0.17, CI 0.05-0.58, *P*<0.001). All five blood samples were tested positive for NS-1 antigen.

**Conclusion:** The presence of accumulated rainwater in pools and empty receptacles around houses acted as breeding grounds for Aedes aegypti mosquitos and was the most probable cause of this outbreak. Following our recommendations, the health department-initiated mosquito breeding sites control activities through residual insecticide spray and advocacy on the use of protective measures against mosquito bites.

## Introduction

Dengue Fever (DF) is a vector-borne illness caused by dengue virus which is transmitted to humans through mosquito Aedes Aegypti and Aedes Albopictus. This mosquito also transmits other infections like chikungunya, yellow fever and zika virus infection. Dengue Virus (DENV) is a single stranded RNA virus that has four serotypes, DEN1, DEN2, DEN3, and DEN4.^1^ Infection with one DENV serotype confers the life time immunity to the individual but reinfection with another serotype can cause a severe infection leading to hemorrhagic fever and shock syndrome. Presence of all four DENV serotypes have been reported in Pakistan with most prevalent serotypes DEN2 and DEN3.^2 3^ Individual with classical DF presents clinically with fever, nausea, vomiting, headache, retro-orbital pain, muscle or joint/bone pain and rash.^4^ Dengue Fever affects both genders, males are predominantly involved.^5^ Epidemics similar to these symptoms have been documented in the past occurred in the 17^th^ century in West Indies and Central America.^6^ Globally about 50% of population is at risk of DF.^7^ Recent estimate indicates 50-100 million cases of dengue fever and 250000-500000 cases of DHF occur every year worldwide.^8^ Asia, Africa and America contribute 70 percent, 16 percent and 14 percent of this burden respectively.^9^ First outbreak of DF in Pakistan was reported in 1994 followed by occurrence of many outbreaks in different cities during last decade.^10^ DF is now widely prevalent and endemic in Pakistan including capital city Islamabad.^11^ Dengue endemic countries have risk of recurrent DF outbreaks.^12^ Major contributing factors include overcrowding/increased population, poor sanitization, unplanned urbanization and international travel and trade.^13^ Monsoon rainy season resulting in floods and accumulation of rain water in pools and empty receptacles also contribute to provide breeding sites for the vector.^14^ Humid and warm environment provide breeding opportunities for the mosquitoes^15^. Most dengue outbreaks are reported during rainy season.^16^ Dengue prevention and control measures include vector control (elimination of mosquito breeding sites, insecticide residual spray and thermal fogging) and prevention of mosquito bites (health education of community).

## Background

Lady health workers reported 14 suspected cases of dengue fever from UC Sohan, Islamabad (a rural UC with a population of 45,747 individuals) to the health department, Islamabad capital territory (ICT) during the second week of October (14^th^ October) 2019. On 17^th^ October 2019, after confirmation of outbreak, on the directive of DHO, FELTP fellow from Federal disease surveillance and response unit (FDSRU) was deputed to conduct the field investigation with the objectives to confirm outbreak, identify possible risk factors and to recommend control measures.

## Methods

Outbreak investigation was conducted from 17^th^ October to 25^th^ November 2019. FELTP-Fellow visited the area and Dengue cases were identified through door-door active case search. A working case definition was developed which was sudden onset of fever (> 38°C) in a resident of union council Sohan, between October 8 to November 25, 2019, with two or more of the following signs/symptoms: headache, retro-orbital pain, arthralgia, myalgia, petechial rash with positive NS1 (Non-structural Protein-1) antigen detection in serum.

After taking an Institutional Review Board permission from The Federal General Hospital Islamabad was obtain vide number: F-3-144/ADMN-EC-FGH. An informed written consent was administered to the literate respondents while a translation was read to the illiterate and guardians/parents of the minors declaring their autonomy, beneficence and justice rendered to them. A pretested, structured questionnaire was used to collect the information on demographics, date of onset of illness, sign/symptoms, risk factors information like use of mosquito coil, repellent lotion, protective full clothing, insecticide sprays, screening of doors, stagnant water around house, presence of water spots inside house (flower pots, open containers with water, water pools, old tires) and travel outside union council 15 days prior to illness and a line list was developed. All identified cases were NS1 positive from different hospitals and laboratories. Blood samples (5ml each) from five suspected cases having fever were collected according to the guidelines set by Public Health laboratory Division, National Institute of health Pakistan and sent for laboratory confirmation. Age and sex matched controls (n=85) were selected for comparison from same area with a 1:1 ratio.

## Results

During the active case search 547 permanent houses and temporary huts (Punjabi Jungian) in UC Sohan were surveyed from 17 October to 25 November, 2019 and total 85 cases (71 cases identified through active case search) of acute dengue fever were identified and interviewed.

The Epi curve showed that the first case had date of onset of illness on 10th October, 2019 and most of the cases developed sign/symptoms on 13th October (n=8), 1^st^ and 4^th^ Nov (n=5) and 2^nd^ and 10^th^ November (n=4) (**Figure1**).

**Figure 1:**
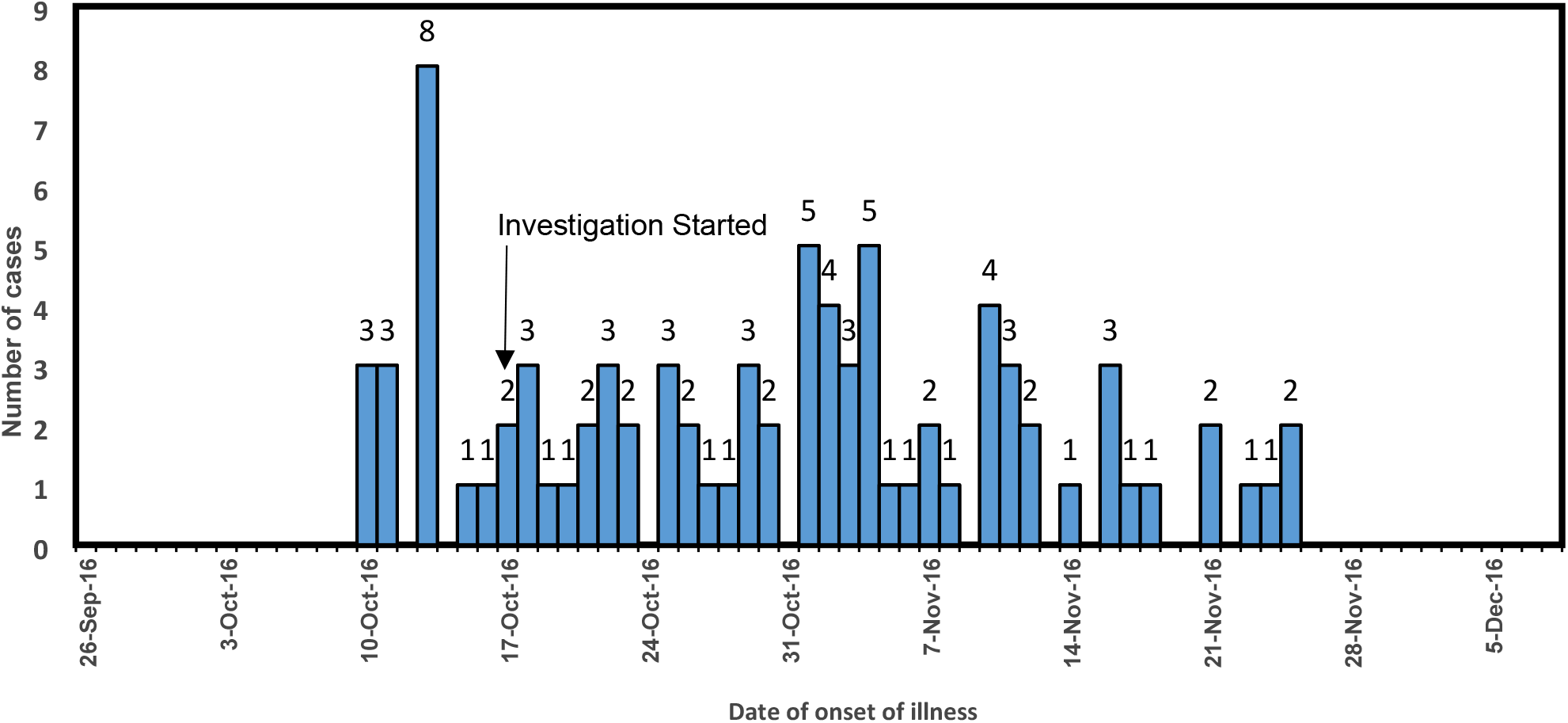
Epi curve – No. of cases by the date of onset of illness, union council Sohan Islamabad October-November 2019

The most frequent sign/symptoms were; Fever 100.00% (n= 85), Headache 83.53% (n=71), Joint/bone pain 75.29% (n= 64), Myalgia 61.18% (n=52), Nausea/Vomiting 49.41% (n=42), Retro-orbital Pain 43.53 % (n= 37) and Abdominal Pain 32.94% (n=28).

Majority (56.5%) of the cases were males (n=48). 35.3% (n=30) of cases were uneducated. Majority 87% (n=74) of the cases said they were using full protective clothing. 81% (n=69) were using mosquito coils/mats in their home. 34% (n=29) cases said they used repellent lotion for mosquito bite prevention. Use of insecticide repellent and door screening is 31% (n=26) and 40% (n=34) respectively. Only 5.8% (n=5) cases used bed nets.

Stagnant water was present around 31.7% (n=27) cases. 65% (n=55) cases had general cleanliness around houses. 4 cases (4.7%) cases said they have old tires on their roof. Presence of water flower pots and open water containers was only2.35% (n=2). None of the cases have water pool inside their home. (**Table 1**).

**Table 1:**
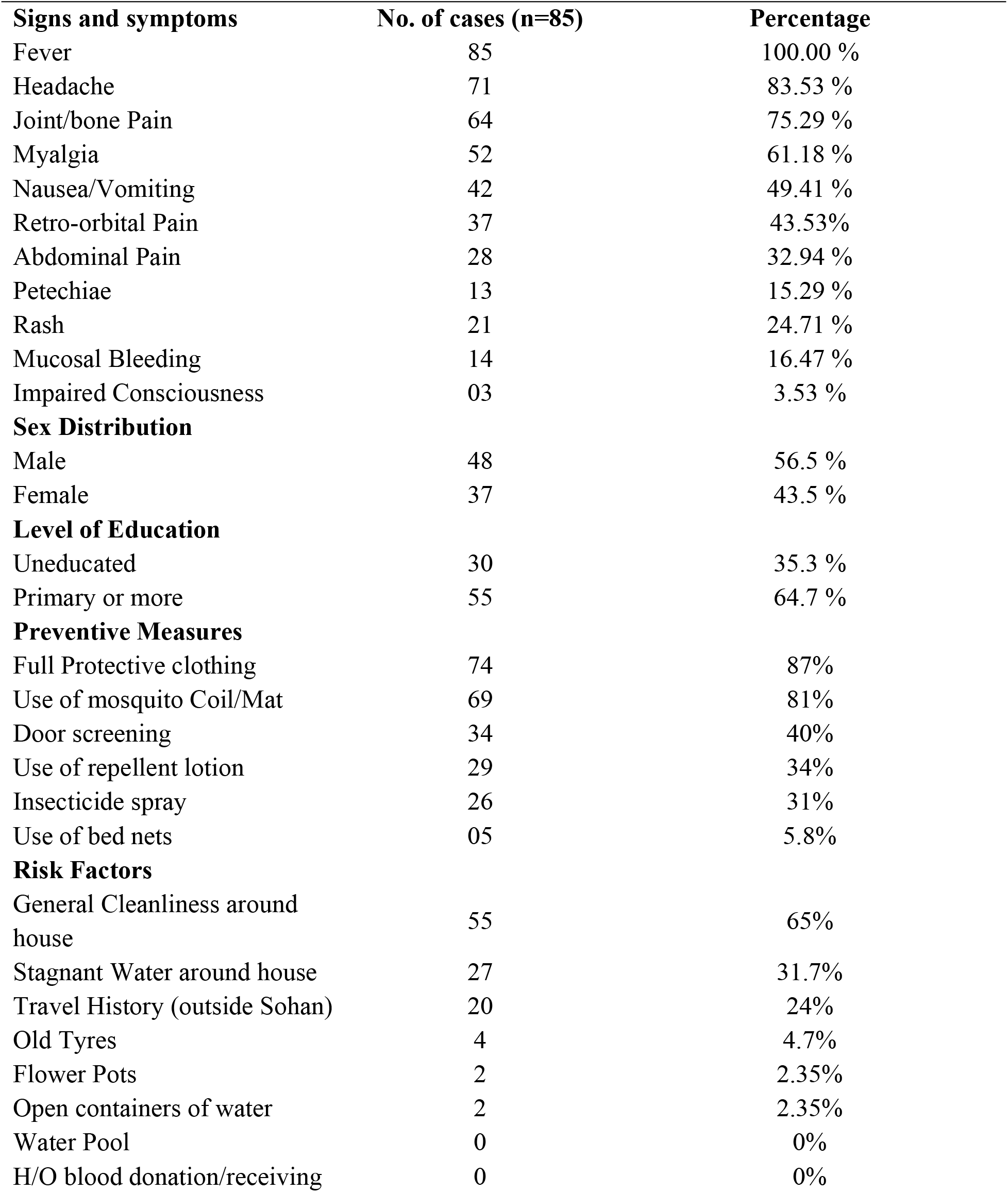
Summary statistics of dengue fever cases, Sohan, Islamabad (n=85) October-November 2019.

| **Signs and symptoms** | **No. of cases (n=85)** | **Percentage** |
| --- | --- | --- |
| Fever | 85 | 100.00 % |
| Headache | 71 | 83.53 % |
| Joint/bone Pain | 64 | 75.29 % |
| Myalgia | 52 | 61.18 % |
| Nausea/Vomiting | 42 | 49.41 % |
| Retro-orbital Pain | 37 | 43.53% |
| Abdominal Pain | 28 | 32.94 % |
| Petechiae | 13 | 15.29 % |
| Rash | 21 | 24.71 % |
| Mucosal Bleeding | 14 | 16.47 % |
| Impaired Consciousness | 03 | 3.53 % |
| **Sex Distribution** |  |  |
| Male | 48 | 56.5 % |
| Female | 37 | 43.5 % |
| **Level of Education** |  |  |
| Uneducated | 30 | 35.3 % |
| Primary or more | 55 | 64.7 % |
| **Preventive Measures** |  |  |
| Full Protective clothing | 74 | 87% |
| Use of mosquito Coil/Mat | 69 | 81% |
| Door screening | 34 | 40% |
| Use of repellent lotion | 29 | 34% |
| Insecticide spray | 26 | 31% |
| Use of bed nets | 05 | 5.8% |
| **Risk Factors** |  |  |
| General Cleanliness around house | 55 | 65% |
| Stagnant Water around house | 27 | 31.7% |
| Travel History (outside Sohan) | 20 | 24% |
| Old Tyres | 4 | 4.7% |
| Flower Pots | 2 | 2.35% |
| Open containers of water | 2 | 2.35% |
| Water Pool | 0 | 0% |
| H/O blood donation/receiving | 0 | 0% |

Mean age of the cases was 34.4 years + 16.05 (range 3-71 years). Most common age group with highest attack rates were 45-54 years (AR=0.43%), 25-34 years (AR=0.34%) and 35-44 years (AR=0.31%). Overall attack rate was 0.19%. (**Table 2)**

**Table 2:**
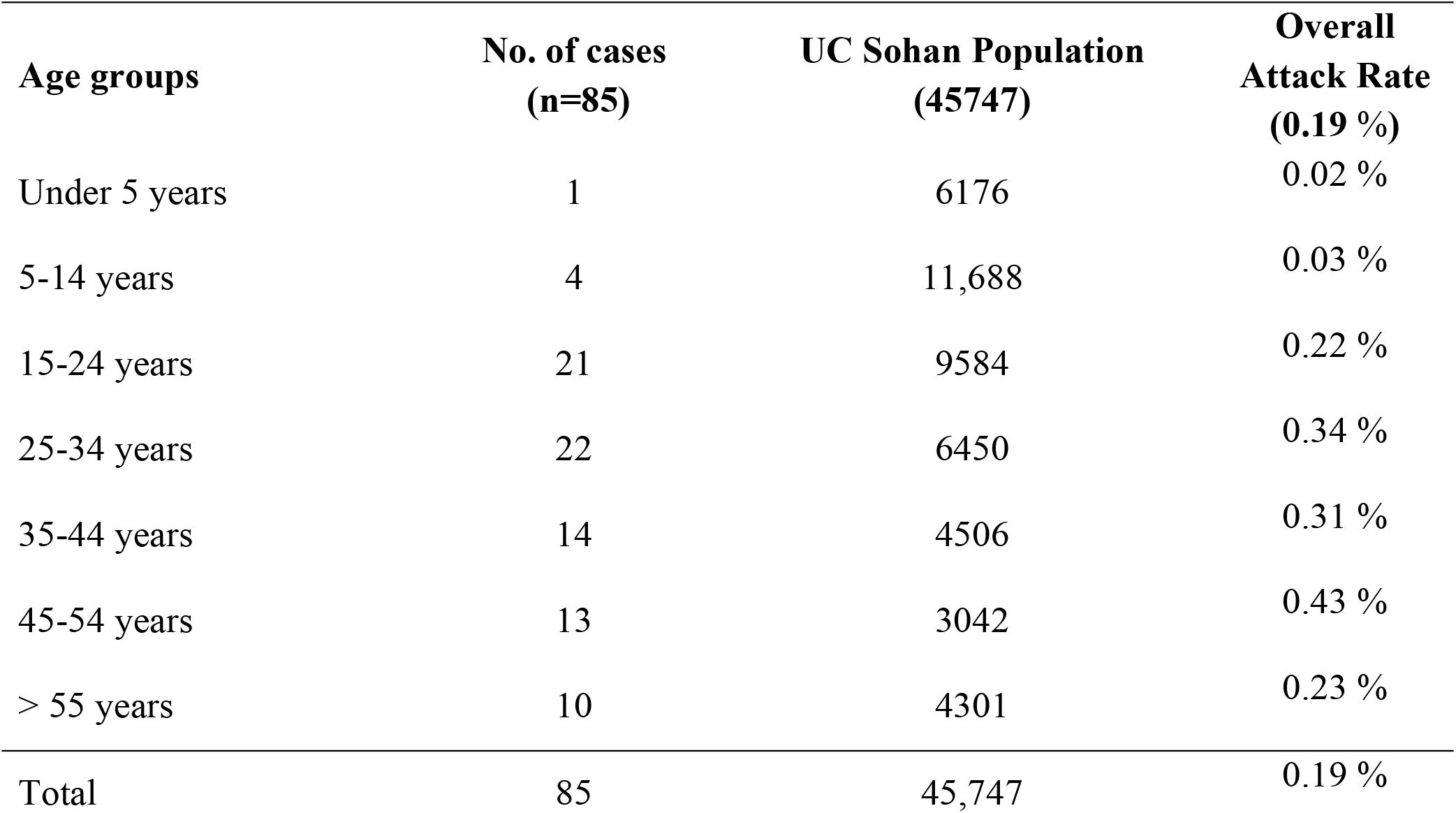
Age specific attack rates of among the dengue fever cases, Sohan Islamabad, October-November 2019.

| **Age groups** | **No. of cases**<br>**(n=85)** | **UC Sohan Population**<br>**(45747)** | **Overall**<br>**Attack Rate**<br>**(0.19 %)** |
| --- | --- | --- | --- |
| Under 5 years | 1 | 6176 | 0.02 % |
| 5-14 years | 4 | 11,688 | 0.03 % |
| 15-24 years | 21 | 9584 | 0.22 % |
| 25-34 years | 22 | 6450 | 0.34 % |
| 35-44 years | 14 | 4506 | 0.31 % |
| 45-54 years | 13 | 3042 | 0.43 % |
| > 55 years | 10 | 4301 | 0.23 % |
| Total | 85 | 45,747 | 0.19 % |

### Epi curve with information on rain and temperature plots

Meteorological investigation showed that rain (99 mm) was observed on 1^st^ September with two showers in the second week of September.

Vector breeding might have started after September 1^st^ rain and the first brood of vector was ready by 10^th^ October. Department of health initiated mosquito breeding site control activities through IRS, thermal fogging and awareness campaign through Lady Health Workers (frontline workers). **(Figure 2)**

**Figure 2:**
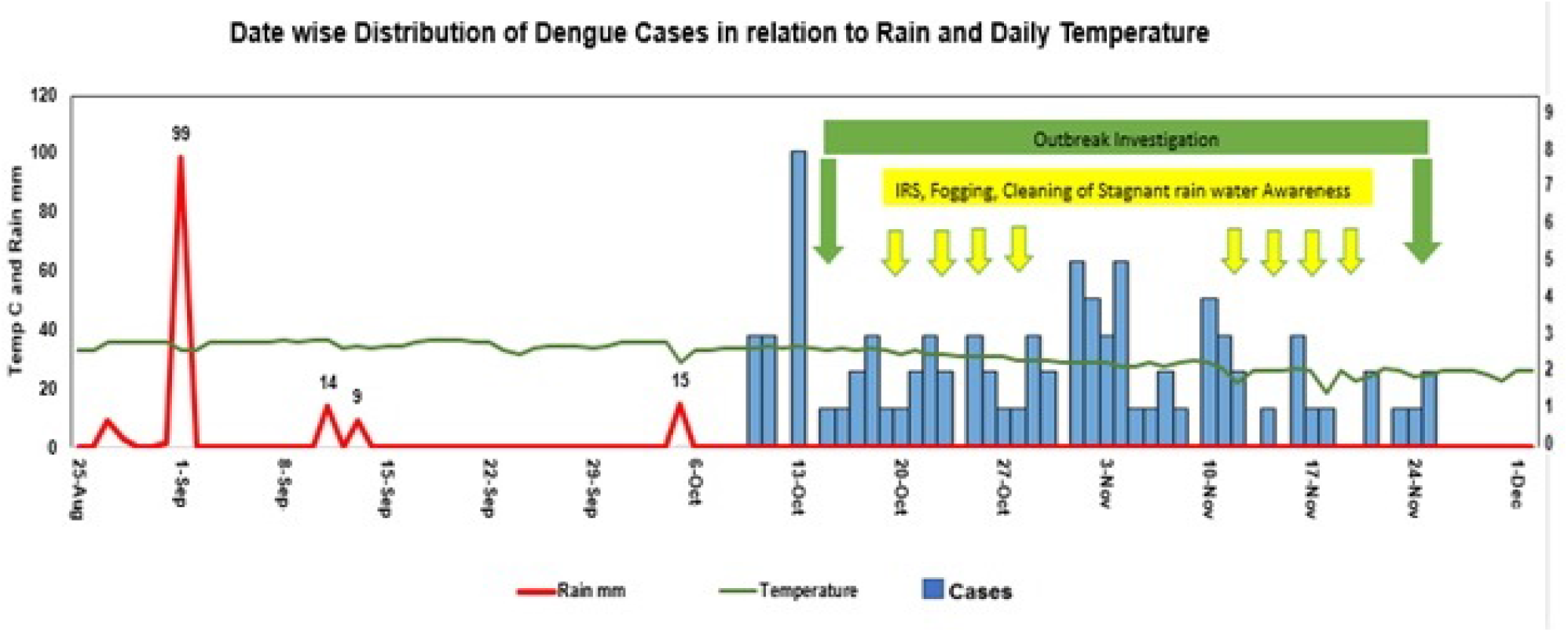
Epi curve with information on rain and temperature plots union council Sohan Islamabad October-November 2019

### Univariate Analysis

Univariate analysis showed the accumulation of rain water around houses (OR 3.90, CI 1.91-7.99, *P*<0.00) is associated with the dengue fever, while use of repellent (OR 0.34, CI 0.18-0.64, *P*<0.00), insecticide spray (OR 0.36, CI 0.19-0.67, *P*<0.001) and full protective clothing (OR 0.13, CI 0.04-0.40, *P*<0.00) has a protective effect. (**Table 3**).

**Table 3:**
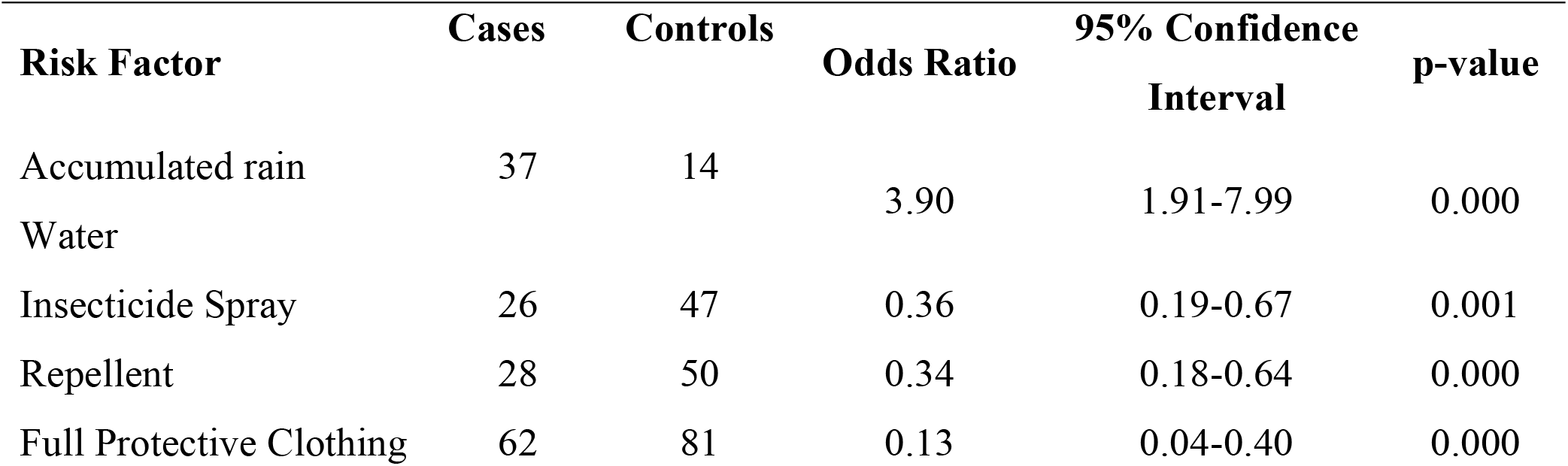
Univariate analysis or risk factors associated with Dengue fever in Islamabad, October-November 2019.

| **Risk Factor** | **Cases** | **Controls** | **Odds Ratio** | **95% Confidence Interval** | **p-value** |
| --- | --- | --- | --- | --- | --- |
| Accumulated rain<br>Water | 37 | 14 | 3.90 | 1.91-7.99 | 0.000 |
| Insecticide Spray | 26 | 47 | 0.36 | 0.19-0.67 | 0.001 |
| Repellent | 28 | 50 | 0.34 | 0.18-0.64 | 0.000 |
| Full Protective Clothing | 62 | 81 | 0.13 | 0.04-0.40 | 0.000 |

### Multivariate Analysis

Multivariate Analysis shows that accumulation of accumulated rain water around houses (OR 2.65, CI 1.2-5.83, *P*<0.01) is associated with the dengue fever, while use of repellent (OR 0.35, CI 0.17-0.70, *P*<0.00), insecticide spray (OR 0.48, CI 0.24-0.97, *P*<0.041) and full protective clothing (OR 0.17, CI 0.05-0.58, *P*<0.00) has a protective effect. (**Table 4**).

**Table 4:** Multivariate analysis of risk factors associated with Dengue fever in Islamabad October-November 2019.

| **Risk Factor** | **Odds Ratio** | **95% Confidence Interval** | **p-value** |
| --- | --- | --- | --- |
| Accumulated rain water | 2.7 | 1.20-5.83 | 0.012 |
| Insecticide spray | 0.5 | 0.24-0.97 | 0.041 |
| Repellent lotion | 0.4 | 0.18-0.70 | 0.003 |
| Full Protective Clothing | 0.2 | 0.05-0.58 | 0.005 |

### Laboratory analysis

Five (05) blood samples taken from acutely ill cases and all were tested positive for dengue fever (NS1 Positive).

## Discussion

Dengue Fever is a significant public health problem which is more common in developing countries. In recent years Pakistan has experienced multiple dengue outbreaks from different geographical areas. This dengue outbreak from union council Sohan was the first ever reported outbreak from this area. Dengue cases were limited to close proximity. Since 1994, several outbreaks have been reported in Pakistan.^17^ Most of the cases reported in this outbreak were males as shown by Khan J.^18^ Males were predominantly involved due to use of non-protective clothing. Most common affected age group with highest attack rates was 45-54 years as shown by Khan J. Most common symptoms were fever and headache followed by joint/bone pains and myalgia. Impaired consciousness was the rarest among all symptoms. There was statistically significant association with accumulated rain water in pools and empty receptacles around houses (p value <0.00) which became breeding sites for mosquito. Insecticide (p value <0.00), repellent (p value <0.00) and full protective clothing (p value <0.00) had a protective effect. This outbreak investigation revealed that protective measures against the vector were not commonly practiced. For the diagnostic purpose NS1 (Non-structural Protein-1) antigen detection test was used as a prime investigation.

## Conclusions

Presence of accumulated rain water in pools and empty receptacles around houses provided breeding site for mosquitos and was the most probable cause of this outbreak. Protective measures against the vector were not commonly practiced. Recommendations were also given regarding Health education of the residents of UC Sohan regarding eliminating the mosquito breeding sites by filling the ponds of accumulated rain water and awareness campaigns for use of repellent, insecticide spray and full protective clothing was recommended. District health department was recommended initiate larvicidal and insecticidal residual spray activities and to strengthen dengue surveillance system.

### Impact of this outbreak investigation

After recommendations, the health department initiated mosquito breeding sites control activities through residual insecticide spray and advocacy on use of protective measures against mosquito bites. Dengue surveillance system was established. No outbreak has been reported since then.

## Data Availability

Data is available with the first author upon request

